# Imprecise assessment of mask use may obscure associations with SARS-CoV-2 positivity

**DOI:** 10.1101/2020.12.30.20249033

**Authors:** Sophie Bérubé, Steven J. Clipman, Shruti H. Mehta, Sunil S. Solomon, Amy Wesolowski

## Abstract

Masks are effective measures to prevent the transmission of SARS-CoV-2, however, lack of a national mandate coupled with poor adherence has led to suboptimal levels of transmission reduction. Although data has suggested that mask adherence is high, few studies have captured details on how mask wearing changes with activities and how these behaviors are associated with SARS-CoV-2 positivity. We recruited an online sample of 3,058 respondents from three US states (MD, FL, IL; n∼1000/state) between September 16 - October 15, 2020. The median age of the sample was 47; 53% were female, 56% were white and 22% were working outside the home. Seventy three percent of the sample reported always wearing a mask indoors and outdoors based on local guidelines, however, 78% of participants who reported always wearing a mask reported taking their mask off when outside the home. While overall masking according to guidelines was not significantly associated with SARS-CoV-2 positivity, sometimes, often or always removing a mask during activities were significantly associated with SARS-CoV-2 positivity (adjusted odds ratio for always vs never removing mask: 9.92; 95% CI: 1.16 – 85.1). These findings suggest that masks were most effective when worn without removal reflecting the need for consistent use.

---

Mask wearing has been shown to be effective at preventing the transmission of SARS-CoV-2 and remains a key public health intervention for mitigating the COVID-19 pandemic (1,2). Mask wearing requires high population-level compliance (4) and is most effective at preventing onward transmission if both infected and susceptible individuals are wearing a mask at the time of contact (3). Inconsistent mask wearing policies and limited enforcement mechanisms have resulted in differential rates of mask uptake across the United States (2–4). Some ecological and individual studies have found associations with reported mask wearing and changes in the SARS-CoV-2 transmission rate or self-reported positivity (1,4,5). To date, the majority of these studies have only asked, general questions about whether or not a person wears a mask when outside the home with options capturing varying degrees of adherence (e.g., always, sometimes, occasionally, and never) (5). A number of other studies in individuals being monitored as part of a contact tracing program have reported that not wearing a mask increases the risk of infection but it is not clear how individuals with a known positive contact may differ from the general population, particularly those without known exposure (6). Accurate, comprehensive information on mask wearing behaviors is critical to inform models that estimate the effectiveness of mask wearing policies along with other non-pharmaceutical interventions.

Using online surveys in three states (Maryland, Florida, and Illinois), we surveyed 3,058 respondents between September 16 - October 15, 2020 to identify patterns in reported mask use by policy and location (indoors vs outdoors), type of activity (while at a bar/restaurant, during fitness activities, and while visiting family/friends), as well as self-reported test positivity within the past two weeks. We also asked individuals questions about other social distancing behaviors and mobility patterns to further capture the differences in possible exposure risks between individuals.

## Materials and Methods

### Survey sample

We recruited participants (September 16 – October 15, 2020) from Florida, Illinois, and Maryland for an online survey about demographics, social distancing practices, and self-reported SARS-CoV-2 PCR positivity in the prior two weeks. States were selected to represent different aspects of the pandemic and differences in state-wide policies. At the time of the survey, only Florida did not have a state-wide mask mandate (7,8). Consenting residents of each state (≥18 years) were recruited using Dynata (https://www.dynata.com), one of the largest first-party global data platforms. Dynata maintains a database of potential participants who are randomized to specific surveys if they meet the demographic targets of the survey. Participants receive modest compensation. Security checks and quality verifications are performed. In order to accrue demographically representative samples, we provided quotas for age, gender, race/ethnicity, and income based on the population composition of the states. Across states, 5,075 were routed to the survey; 714 did not start the survey, 694 started but did not complete the survey, and 609 responses were excluded for non-eligibility.

### Mask usage and activity measures

We asked individuals to respond with ‘always’, ‘sometimes’, ‘never’, or ‘prefer not to say’ if they wear a mask according to local/state guidelines when indoors, outdoors, and when within 6 feet of another individual indoors and outdoors. We further asked participants whether they removed their mask while indoors or outdoors at another individuals’ home, in a bar or restaurant, or while working out at a gym or part of a group fitness class. Individuals could respond with ‘never’, ‘sometimes’, or ‘always’ to these questions. Individuals also responded to questions about their behaviors: if they had visited another individual’s home, a bar, a restaurant, and a gym/outdoor fitness class indoors and outdoors. An individual’s activity score was computed by assigning a person one point for each of the six possible activities they reported participated in (attending an indoor or outdoor bar/restaurant, visiting with friends, relatives or neighbors indoors or outdoors, and attending an indoor gym or outdoor group fitness activity). The sum of points across all activities was the activity score (range: 0 to 6).

To create a mask removal score, for each activity an individual participated in, we assigned zero points for never taking of their mask, one point for sometimes taking off their mask, and two points for always taking off their mask. A person’s overall mask wearing score was the sum of their points, across all the activities they reported, the maximum number of activities was 6 making the maximum mask score 12. We further divided individuals into four mask score categories in accordance with the observed quartiles of the population’s mask wearing scores (quartile 1: 0-4, quartile 2: 4-6, quartile 3: 6-8, quartile 4: 8-12).

### Statistical Methods

Statistical analyses were carried out using R (v3.5.1). Logistic regression was used to analyze associations with SARS-CoV-2 PCR positivity. All variables were used in both univariable and multivariable analyses.

## Results

Individuals in the sample had a median age 47, 53% identified as female, 56% identified as White or Caucasian, 14% as Black or African American, 6% as Hispanic or Latinx, 21% as Asian or Pacific Islander and 3% as American Indian or Alaskan Native. The majority, 53%, had completed a bachelor’s degree or graduate degree and 39% reported working from home while 22% working outside the home. These patterns were consistent across the three states (Table S1).

The majority of individuals reported always wearing a mask indoors (73%) or outdoors (73%) according to state and local policies and this did not differ in a statistically significant way by state (Table S2). In both instances, only approximately 10% reported never wearing a mask indoors or outdoors and only 5% of the sample reported never wearing a mask in both locations (indoors and outdoors). It was more common to report never wearing a mask outdoors, but wearing one sometimes or always indoors (7%) than the reverse (4%).

Younger individuals and those who had no schooling beyond a high school diploma or its equivalent had a significantly increased odds of never wearing a mask indoors or outdoors but we did not observe significant differences by gender, or race/ethnicity in the odds of never wearing a mask indoors or outdoors (Table S3). Of individuals who reported always wearing a mask indoors (n=2047) or outdoors (n=2094), most also reported always wearing a mask when within 6 feet of someone else (indoors: 90%; outdoors: 91%). There were substantially fewer individuals who reported only sometimes wearing a mask indoors (n=498) or outdoors (n=581) and a much smaller percentage of these individuals reported always wearing a mask when within 6 feet of someone else both indoors (29%) and outdoors (32%).

Although overall mask use was high based on these metrics, the majority of individuals also reported taking off their mask when visiting family/friends in another’s home, at a bar or restaurant, or while participating in a group fitness/attending a gym. Of those individuals who visited an indoor (n=935) or outdoor (n=780) bar or restaurant, more than 50% reported always taking their mask off to eat or drink (55% (indoor), 52% (outdoor)). Individuals were more likely to always take off their mask while participating in outdoor fitness activities (57%) than indoor fitness/gym (46%, 149/324). These patterns were consistent, even for those who reported always wearing a mask indoors and outdoors, specifically, a majority of these individuals reported always taking off their masks at a bar or restaurant (indoors: 53%; outdoors: 52%) and many of these people also reported taking off their mask while doing gym/fitness activities (indoors: 44%, outdoors: 64%) The proportion of people removing their mask during activities increased with activity score but was consistently over 50% among those who reported sometimes or always wearing their mask and across all activity scores (Figure 1).

**Figure 1:**
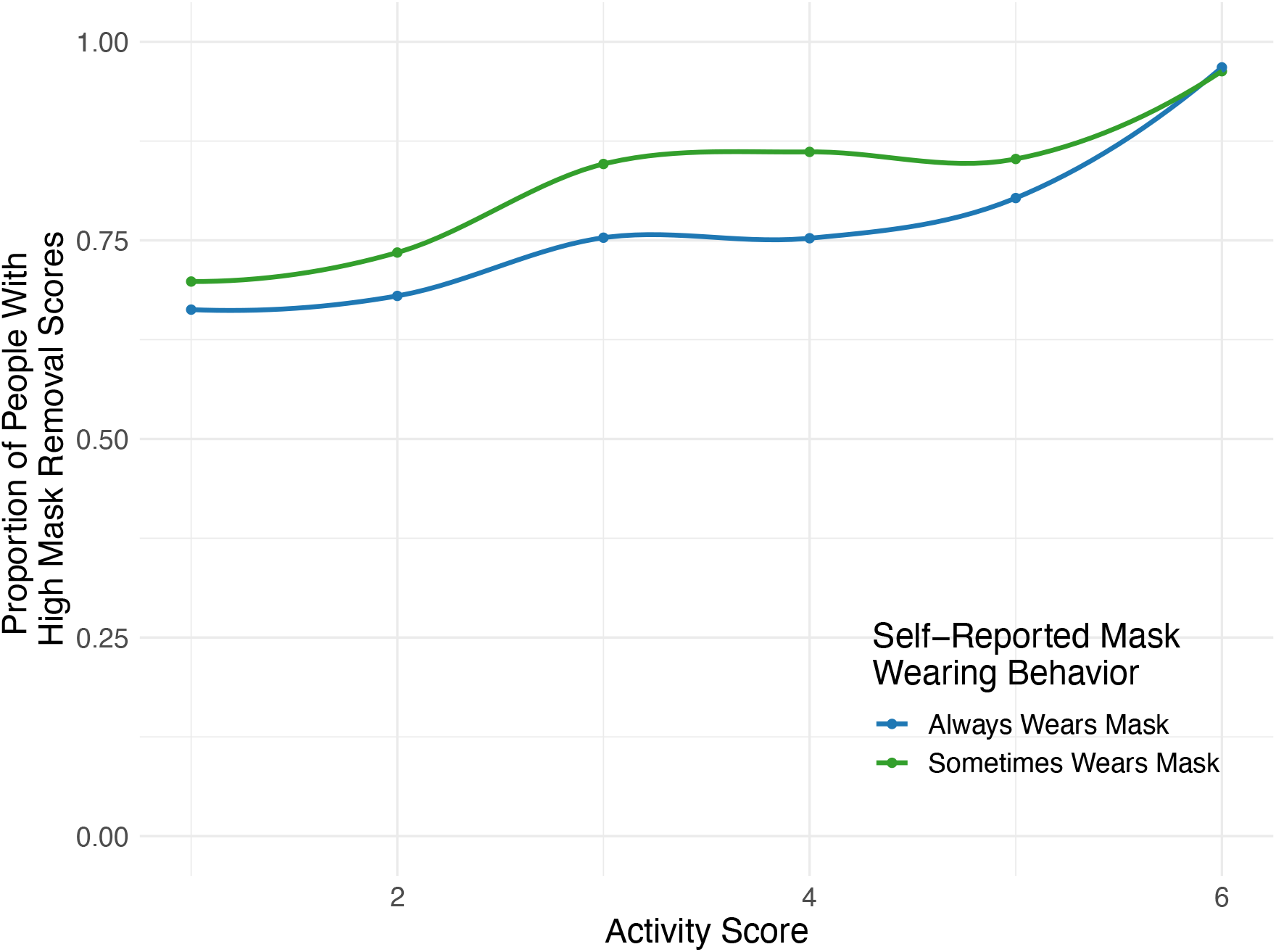
Mask Removal Scores by Level of Activity and Self-Reported Mask Wearing Behavior. Individuals who reported always (blue) or sometimes (green) wearing a mask to the general self-reported mask wearing behavior question (based on local or state guidance) by activity score (higher scores = more activities). For individuals within an activity score, the proportion of people who have a high mask removal score (greater than 4). Higher values indicate they are more report removing their mask for certain activities.

We further investigated the relationship between mask wearing and self-reported SARS-CoV-2 PCR positivity within the past two weeks. Of the people who received a test in the past two weeks, 23% were positive with fairly consistent trends across states (Table S1). Visiting more locations increased the odds of being positive (OR = 2.09; 95% CI: 1.68 – 2.59, Table S4). Adjusted for the number of locations a person visited and other demographic variables, a model that included the masking score as a covariate revealed a statistically significantly increased odds of PCR positivity in the past two weeks for those who removed their mask more often (aOR = 9.92; 95% CI: 1.16-85.1 for always vs never removing mask, Table 1). However, a model that included overall self-reported mask wearing indoors and outdoors as covariates instead, did not reveal a statistically significantly different odds of being PCR positive in the last 2 weeks (never versus always wearing indoors: aOR = 0.81; 95% CI: 0.17-3.39, outdoors: aOR = 1.05; 95% CI: 0.26-3.88) (Table 1). Unadjusted odds ratios are shown in Table S4. Finally, a model including those who did not receive a PCR test in the past two weeks as negative revealed similar results, although the association between removing one’s mask and the odds of being PCR positive in the past two weeks did not rise to the level of significance (Table S5).

**Table 1:**
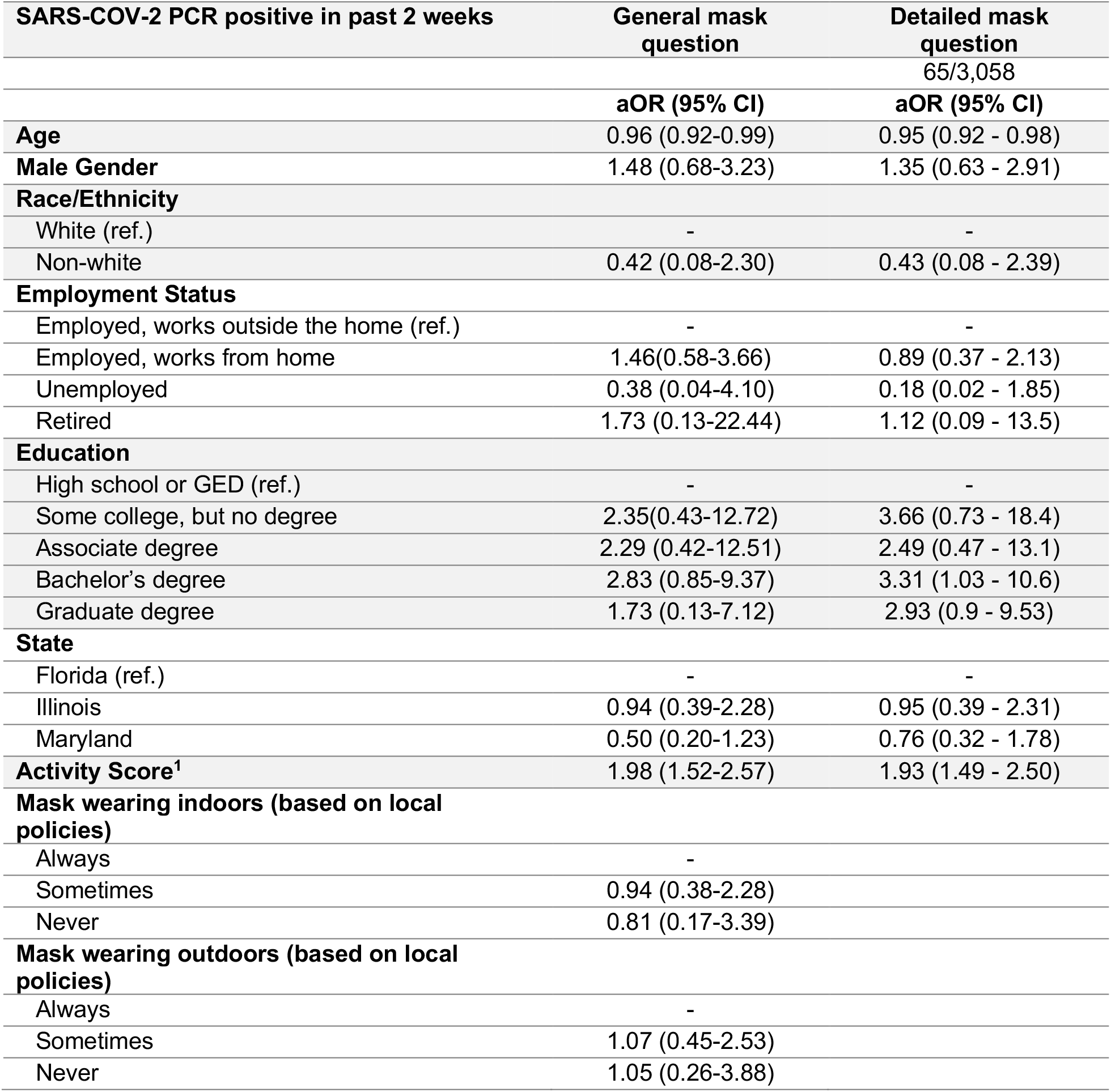
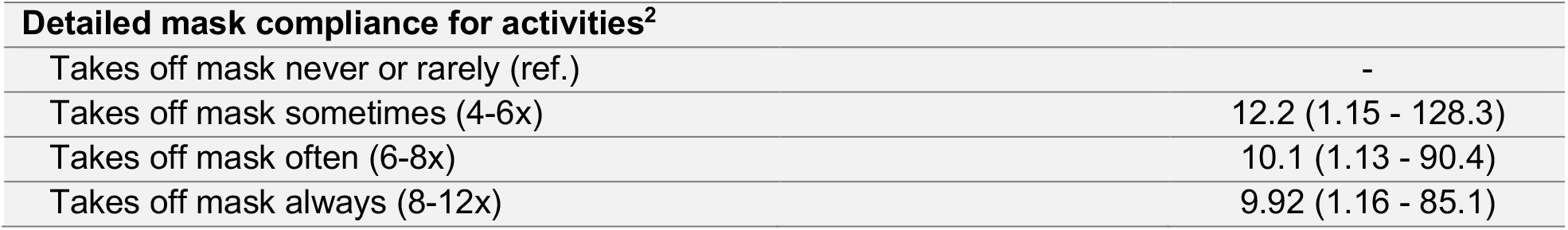
Factors associated with self-reported SARS-CoV-2 PCR positivity in the prior 2 weeks. For those who were tested, a logistic regression model was used to assess the relationship between demographic, activity scores, and mask wearing variables with SARS-CoV-2 positivity. Two models including general mask questions (did you wear a mask indoors or outdoors based on local policies) and detailed mask questions (did you take off your mask during certain activities) were run.

| **SARS-COV-2 PCR positive in past 2 weeks** | **General mask question** | **Detailed mask question** |
| --- | --- | --- |
|  |  | 65/3,058 |
|  | **aOR (95% CI)** | **aOR (95% CI)** |
| **Age** | 0.96 (0.92-0.99) | 0.95 (0.92 - 0.98) |
| **Male Gender** | 1.48 (0.68-3.23) | 1.35 (0.63 - 2.91) |
| **Race/Ethnicity** |  |  |
| White (ref.) | - | - |
| Non-white | 0.42 (0.08-2.30) | 0.43 (0.08 - 2.39) |
| **Employment Status** |  |  |
| Employed, works outside the home (ref.) | - | - |
| Employed, works from home | 1.46(0.58-3.66) | 0.89 (0.37 - 2.13) |
| Unemployed | 0.38 (0.04-4.10) | 0.18 (0.02 - 1.85) |
| Retired | 1.73 (0.13-22.44) | 1.12 (0.09 - 13.5) |
| **Education** |  |  |
| High school or GED (ref.) | - | - |
| Some college, but no degree | 2.35(0.43-12.72) | 3.66 (0.73 - 18.4) |
| Associate degree | 2.29 (0.42-12.51) | 2.49 (0.47 - 13.1) |
| Bachelor's degree | 2.83 (0.85-9.37) | 3.31 (1.03 - 10.6) |
| Graduate degree | 1.73 (0.13-7.12) | 2.93 (0.9 - 9.53) |
| **State** |  |  |
| Florida (ref.) | - | - |
| Illinois | 0.94 (0.39-2.28) | 0.95 (0.39 - 2.31) |
| Maryland | 0.50 (0.20-1.23) | 0.76 (0.32 - 1.78) |
| **Activity Score<sup>1</sup>** | 1.98 (1.52-2.57) | 1.93 (1.49 - 2.50) |
| **Mask wearing indoors (based on local policies)** |  |  |
| Always | - |  |
| Sometimes | 0.94 (0.38-2.28) |  |
| Never | 0.81 (0.17-3.39) |  |
| **Mask wearing outdoors (based on local policies)** |  |  |
| Always | - |  |
| Sometimes | 1.07 (0.45-2.53) |  |
| Never | 1.05 (0.26-3.88) |  |

<sup>1</sup> An individual's activity score is computed by assigning a person one point for each activity they reported participating in. There were six possible activities contributing to the score: attending an indoor or outdoor bar/restaurant, visiting with friends, relatives or neighbors indoors or outdoors, and attending an indoor gym or outdoor group fitness activity. The sum of points across all activities is the activity score, scores ranged from 0 to 6.

| **Detailed mask compliance for activities<sup>2</sup>** |  |
| --- | --- |
| Takes off mask never or rarely (ref.) | - |
| Takes off mask sometimes (4-6x) | 12.2 (1.15 - 128.3) |
| Takes off mask often (6-8x) | 10.1 (1.13 - 90.4) |
| Takes off mask always (8-12x) | 9.92 (1.16 - 85.1) |

## Discussion

In this sample of individuals across three US states, we demonstrated that although the majority of individuals reported always wearing a mask indoors and outdoors based on local guidelines and when within 6 feet of another individual, more than half of these individuals also reported removing their masks while participating in activities outside the home.

The general question about overall mask wearing in accordance with state and local guidelines was not associated with SARS-CoV-2 positivity within the past two weeks when adjusted for the individual’s activity score and other demographic variables. However, when nuanced information about mask wearing behavior during specific activities was incorporated, individuals who wore a mask less frequently were significantly more likely to be positive for SARS-CoV-2 in the last two weeks. These results suggest that additional detail about mask wearing behaviors, beyond general questions that have commonly been used in studies to date, can provide a measurement that is able to more accurately capture risk of infection as a function of mask wearing and measure the population’s adoption of mask use (5).

Unlike other studies, we did not see a large difference in demographics (age, education level, employment status, gender and race) among those who reported always wearing a mask compared to those who reported sometimes or never wearing one, however, age and education level were significantly associated (9,10). In addition, these questions and data remain imperfect; there are many facets of mask-wearing behavior that the survey questions were unable to capture. For example, these questions did not consider how long people took off or kept on their mask while outside of their homes nor did we ask questions about wearing a mask below the nose or the type of mask worn. Further, online surveys require internet access and may bias population samples. However, internet access is fairly high in the locations where we conducted our survey (11). It is likely that these online surveys miss individuals with lower levels of education and homeless persons, two groups where mask may be lower; therefore, these estimates may be higher than what might be expected in the population. While we worked with the survey company to balance responses to reflect the age, income, race/ethnicity, and gender of the adult populations of each state, this process was imperfect. In addition, we asked about self-reported SARS-CoV-2 positivity which does not capture differential access to testing although we performed sensitivity analysis by including and removing those who did not receive a test and found similar associations although not to the level of statistical significance.

Adherence to public health recommendations is never perfect but to date, few studies have related mask wearing and the nuances in those behaviors with infection on the individual level. These results suggest that more granular questions about mask wearing behavior in particular in relation to specific activities are critical to calibration of models. Messaging on the need for consistent and correct mask use are urgently needed to curb transmission.

## Data Availability

Data to support these findings are available from the corresponding author upon reasonable request.

## Acknowledgements

We would like to gratefully acknowledge Mr. Adebola Adegbesan who worked closely with our team in the recruitment of the study sample.

## Footnotes

1 An individual’s activity score is computed by assigning a person one point for each activity they reported participating in. There were six possible activities contributing to the score: attending an indoor or outdoor bar/restaurant, visiting with friends, relatives or neighbors indoors or outdoors, and attending an indoor gym or outdoor group fitness activity. The sum of points across all activities is the activity score, scores ranged from 0 to 6.

2 A person’s overall mask wearing score is the sum of their points, across all the activities they reported. In order to obtain the mask wearing variable in Tables 1 and S5, we divided individuals into four categories based on their mask wearing score in accordance with the observed quartiles of the population’s mask wearing scores (quartile 1: 0-4, quartile 2: 4-6, quartile 3:6-8, quartile 4: 8-12).

