## Supplementary Information for "Imprecise assessment of mask use may obscure associations with SARS-CoV-2 positivity"

**Table S1.** Characteristics of overall study sample by state.

| | Total | FL | IL | MD |
|---|---|---|---|---|
| | n = 3,058 | n = 998 | n = 1,045 | n = 1,015 |
| **Median Age (IQR)** | 47 (33 – 64) | 48 (33 – 65) | 47 (35 – 64) | 46 (32 – 63) |
| **Median Household Size (IQR)** | 2 (2 – 3) | 2 (2 – 3) | 2 (1 – 3) | 2 (2 – 3) |
| **Gender, n (%)** | | | | |
| Female | 1,612 (53%) | 537 (51%) | 545 (52%) | 477 (53%) |
| Male | 1,431 (47%) | 505 (49%) | 502 (48%) | 422 (47%) |
| Other | 1 (0.03%) | 0 (0%) | 0 (0%) | 1 (0.1%) |
| **Race/Ethnicity, n (%)** | | | | |
| White/Caucasian | 1,712 (56%) | 444 (45%) | 628 (61%) | 640 (64%) |
| Black/African American | 632 (21%) | 223 (22%) | 169 (16%) | 240 (24%) |
| Hispanic/Latino | 437 (14%) | 241 (24%) | 141 (14%) | 55 (5%) |
| Asian/Pacific Islander | 175 (6%) | 58 (6%) | 65 (6%) | 52 (5%) |
| Other | 75 (3%) | 25 (3%) | 33 (3%) | 17 (2%) |
| **Educational Attainment, n (%)** | | | | |
| High school degree or less | 552 (18%) | 181 (18%) | 192 (18%) | 179 (18%) |
| Associate degree | 544 (18%) | 167 (17%) | 207 (20%) | 170 (17%) |
| Some college (no degree) | 335 (11%) | 133 (13%) | 122 (12%) | 80 (8%) |
| Bachelor's degree | 935 (31%) | 299 (30%) | 320 (31%) | 316 (31%) |
| Graduate degree | 672 (22%) | 213 (22%) | 198 (19%) | 261 (26%) |
| **Annual Household Income, n (%)** | | | | |
| < $20,000 | 336 (11%) | 134 (13%) | 101 (10%) | 101 (10%) |
| $20,000 – $39,000 | 481 (16%) | 194 (19%) | 169 (16%) | 118 (12%) |
| $40,000 – $49,000 | 250 (8%) | 103 (10%) | 82 (8%) | 65 (6%) |
| $50,000 – $69,000 | 548 (18%) | 171 (17%) | 215 (21%) | 162 (16%) |
| $70,000+ | 1,443 (47%) | 396 (40%) | 478 (46%) | 569 (56%) |
| **Employment Status, n (%)** | | | | |
| Employed, working outside the home | 1,168 (37%) | 361 (37%) | 427 (41%) | 380 (38%) |
| Employed, working from home | 878 (29%) | 275 (28%) | 299 (29%) | 304 (30%) |
| Unemployed | 309 (10%) | 102 (10%) | 110 (11%) | 97 (10%) |
| Retired | 664 (22%) | 245 (25%) | 199 (19%) | 220 (22%) |
| **Urban-Rural Classification** | | | | |
| Urban | 1,018 (33%) | 433 (43%) | 486 (47%) | 99 (10%) |
| Suburban | 1,764 (58%) | 511 (51%) | 385 (37%) | 868 (86%) |
| Rural | 276 (9%) | 54 (5%) | 174 (17%) | 48 (5%) |
| **Reported Exposure, n (%)** | 235 (8%) | 98 (10%) | 79 (8%) | 58 (6%) |
| **Reported Symptoms, n (%)** | 223 (7%) | 92 (9%) | 71 (7%) | 60 (6%) |
| **Reported Travel for Any Purpose, n (%)** | 494 (16%) | 184 (18%) | 142 (14%) | 168 (17%) |
| **Received PCR Test in the Last 2 Weeks n (%)** | 281 (9%) | 117(12%) | 80(8%) | 84(8%) |
| **PCR Positive in the Last 2 Weeks, n (%)** | 65 (23%) | 34 (29%) | 15 (19%) | 16 (19%) |

Note: Numbers may not sum to the total if there were participants who elected not to answer a given question, or participants for whom the question was not applicable.

**Table S2:** Proportions of people who always report wearing a mask indoors, outdoors and in both locations by state.

| | Total | FL | IL | MD |
|---|---|---|---|---|
| | **n = 2,849** | **n = 937** | **n = 972** | **n = 940** |
| **Proportion Who Always Wear a Mask Indoors** | 2,087 (73%) | 681 (73%) | 695 (72%) | 708 (75%) |
| **Proportion Who Always Wear a Mask Outdoors** | 2,099 (74%) | 692 (74%) | 691 (71%) | 716 (76%) |
| **Proportion Who Always Wear a Mask Indoors and Outdoors** | 1,682 (59%) | 560 (60%) | 548 (56%) | 574 (61%) |
| **Proportion Who Remove Mask At a Bar or Restaurant** | 2,555(90%) | 826 (88%) | 874 (90%) | 855 (91%) |
| **Proportion Who Remove Mask While Exercising** | 2,212 (78%) | 739 (79%) | 744 (77%) | 729 (78%) |

**Table S3:** Factors associated with never wearing a mask indoors or outdoors: results from unadjusted logistic regression analysis (n=3058)

| | Never wearing a mask indoors or outdoors |
|---|---|
| | **OR (95% CI)** |
| **Age** | 0.98 (0.98 - 0.998) |
| **Male Gender** | 0.86 (0.61 - 1.22) |
| **Race/Ethnicity** | |
| White | - |
| Not white | 0.51 (0.23 - 1.33) |
| **Education** | |
| High school or GED | - |
| Some college, college degree, associates degree or graduate degree | 2.03 (1.38 - 2.94) |

**Table S4:** Factors associated with self-reported PCR test positivity: results from univariate logistic regression analyses (n=218). The indoor and outdoor mask variables are self-reported adherence to local policies. Participants were asked whether they wore a mask indoors and outdoors in accordance with local guidelines, the options for answering were never, always sometimes for each location.

| | **SARS-CoV-2 PCR positive in the past 2 weeks** |
|---|---|
| | **OR (95% CI)** |
| **Age** | 0.96 (0.94 - 0.99) |
| **Male Gender** | 1.65 (0.93 - 2.91) |
| **Race/Ethnicity** | |
| White (ref.) | - |
| Non-white | 0.59 (0.14 - 2.43) |
| **Employment Status** | |
| Employed, works outside the home (ref.) | - |
| Employed, works from home | 1.65 (0.82 - 3.36) |
| Unemployed | 0.35 (0.04 - 2.99) |
| Retired | 0.21 (0.03 - 1.74) |
| **Education** | |
| High school or GED (ref.) | - |
| Some college, but no degree | 0.78 (0.23 - 2.71) |
| Associate degree | 0.83 (0.22 - 3.16) |
| Bachelor's degree | 1.79 (0.7 - 4.53) |
| Graduate degree | 2.13 (0.83 - 5.49) |
| **State** | |
| Florida (ref.) | - |
| Illinois | 0.56 (0.28 - 1.12) |
| Maryland | 0.57 (0.29 - 1.13) |
| **Activity Score** | 2.09 (1.68 - 2.59) |
| **Masking wearing indoors (based on policies)** | |
| Always | - |
| Sometimes | 1.08 (0.57-2.04) |
| Never | 0.96 (0.36-2.54) |
| **Masking wearing outdoors (based on policies)** | |
| Always | - |
| Sometimes | 1.01 (0.51-2.02) |
| Never | 0.87 (0.28-2.74) |
| **Detailed mask compliance score for activities** | |
| Takes off mask never or rarely (ref.) | - |
| Takes off mask sometimes | 7.20 (0.82 - 63.2) |
| Takes off mask often | 9.60 (1.22 - 75.5) |
| Takes off mask always | 16.8 (2.23 - 126.7) |

**Table S5:** Factors associated with self-reported SARS-CoV-2 PCR positivity in the prior 2 weeks among the full study sample (n=3,058). Analysis compares those who were PCR positive in the prior 2 weeks (n=65) to those who tested negative in the prior 2 weeks and those who were not tested.

| | **SARS-CoV-2 PCR positive in the past 2 weeks** |
|---|---|
| | **aOR (95% CI)** |
| **Age** | 0.95 (0.92 - 0.97) |
| **Male Gender** | 1.40 (0.76 - 2.62) |
| **Race/Ethnicity** | |
| White (Ref.) | - |
| Not white | 0.48 (0.14 - 2.32) |
| **Employment Status** | |
| Employed, works outside the home (Ref.) | - |
| Employed, works from home | 1.28 (0.64 - 2.73) |
| Unemployed | 0.25 (0.01 - 1.41) |
| Retired | 0.55 (0.03 - 3.70) |
| **Education** | |
| High school or GED (Ref.) | - |
| Some college, but no degree | 1.60 (0.42 - 5.89) |
| Associate degree | 1.59 (0.36 - 6.25) |
| Bachelor's degree | 2.89 (1.14 - 8.30) |
| Graduate degree | 2.23 (0.84 - 6.55) |
| **State** | |
| Florida (Ref.) | - |
| Illinois | 0.60 (0.28 - 1.20) |
| Maryland | 0.65 (0.32 -1.29) |
| **Masking Score** | |
| Takes off mask never or rarely (Ref.) | - |
| Takes off mask sometimes | 5.19 (0.82 - 101.3) |
| Takes off mask often | 6.74 (1.26 - 125.2) |
| Takes off mask always | 7.34 (1.46 - 133.7) |
| **Activity Score** | 2.56 (2.07 - 3.24) |
